# Islet autoantibody level distributions in type 1 diabetes and their association with genetic and clinical characteristics

**DOI:** 10.1101/2021.08.04.21261472

**Authors:** Sian Louise Grace, Jack Bowden, Helen C. Walkey, Akaal Kaur, Shivani Misra, Beverley M. Shields, Trevelyan J. McKinley, Nick S Oliver, Timothy McDonald, Desmond G. Johnston, Angus G. Jones, Kashyap Amratial Patel

**Affiliations:** The Institute of Biomedical & Clinical Science, University of Exeter Medical School, Exeter, U.K.; Division of Diabetes, Endocrinology and Metabolism, Imperial College London, London, U.K.; Academic Department of Clinical Biochemistry, Royal Devon and Exeter NHS Foundation Trust, Exeter, U.K.; Macleod Diabetes and Endocrine Centre, Royal Devon and Exeter NHS Foundation Trust, Exeter, U.K.

## Abstract

Positivity for islet autoantibodies is used for diagnosis of type 1 diabetes. However, the importance of the autoantibody level at diagnosis of type 1 diabetes is not clear. Here, we assessed the association of glutamate decarboxylase (GADA), islet antigen-2 (IA-2A) and zinc transporter 8 (ZnT8A) autoantibody levels, measured using radiobinding assays, on genetic and clinical characteristics at diagnosis of 1536 participants with diabetes who were positive for these autoantibodies. We show that GADA and IA-2A levels had bimodal distributions, but ZnT8A level did not. The comparison of genetic and clinical characteristics between high and low level categories showed high GADA level was associated with older age at diagnosis, female sex and *HLA-DR3-DQ2*, whereas high IA-2A level was associated with younger age of diagnosis, ZnT8A positivity and *HLA-DR4-DQ8*. We replicated our findings in an independent cohort of 427 people with type 1 diabetes where autoantibodies were measured using enzyme-linked immunosorbent assays. In conclusion, Islet autoantibody levels provide additional information over positivity in type 1 diabetes at diagnosis. The bimodality of islet autoantibody levels highlights the novel aspect of heterogeneity of type 1 diabetes which may have implications on prediction, treatment and prognosis.

Islet autoantibodies are commonly used in the diagnosis and prediction of type 1 diabetes. They are well established as the biomarkers of the underlying autoimmune pathogenesis (1). Autoantibodies to islet cell antigen (ICA), glutamate decarboxylase (GADA), islet antigen-2 (IA-2A), insulin (IAA) and zinc transporter 8 (ZnT8A) are the most commonly used islet autoantibodies at diagnosis (2). As detectable islet autoantibodies overlap between health and disease, a test is usually considered positive for a given islet autoantibody when the antibody level is higher than a 97.5–99^th^ centile of a control population (3; 4). In routine clinical practice, quantitative islet autoantibody results are usually interpreted as positive or negative, and the level of the islet autoantibody, is not thought to be clinically meaningful.

Islet autoantibody levels may provide additional information over positivity in type 1 diabetes at diagnosis. Similar to type 1 diabetes, autoantibodies to a specific antigen are commonly used for diagnosis in many other autoimmune diseases (such as TSH receptor antibodies in Graves’ disease and Rheumatoid Factor and Citrullinated Protein in rheumatoid arthritis). For Graves’ disease and rheumatoid arthritis, along with autoantibody positivity for these antigens, autoantibody level at diagnosis is associated with disease severity, prognosis and treatment success (5; 6). Multiple studies have shown a role for islet autoantibody level in the prediction of onset of type 1 diabetes, those with a higher levels of IA-2A, IAA and ICA have an increased risk of developing type 1 diabetes in at-risk populations (1; 7-9). However, it is not clear if the islet autoantibody level at diagnosis of type 1 diabetes, in addition to its interpretation as ‘positive’, is associated with the clinical phenotype similar to other autoimmune diseases.

In this study, we undertook an analysis of GADA, IA-2A and ZnT8A levels at diagnosis in a large cohort of participants with type 1 diabetes, assessing the association of islet autoantibody levels on genetic and clinical characteristics at diagnosis in people with type 1 diabetes.

## Research Design and Methods

### Study cohorts

We recruited 1536 participants with a clinician-assigned diagnosis of type 1 diabetes (age at diagnosis range 4–74 years) who were positive for any of GADA, IA-2A or ZnT8A at diagnosis. These participants were recruited as part of the UK-wide ADDRESS-2 study. The detailed protocol for the study has been published previously (10). The included participants were recruited at diagnosis (<6 months), were >4y of age, insulin-treated from diagnosis, and were self-reported to be of white European ethnicity. We excluded individuals with non-European ethnicity as the type 1 diabetes genetic risk score is only validated for those with European ancestry. DNA and serum samples, clinical data (including the characteristics of diabetes at diagnosis) were collected at recruitment. The overall cohort characteristics are provided in ESM Table 1.

**Table 1:** Comparison of clinical characteristics at diagnosis between high and low GADA level groups for GADA positive type 1 diabetes cases. Bimodal GADA level distribution was divided into two groups using the nadir between the two modes at 452 DK U/ml. Values expressed as median (interquartile range) unless stated. ^#^ Only for people who were positive for that antibody * indicates a p value lower than threshold the *P* value for multiple comparisons (0.05/19 =0.0026)

| Characteristic | Low Level GADA | High Level GADA | P value |
| --- | --- | --- | --- |
| n (%) | 711 (56) | 557 (44) |  |
| GADA level (DK U/ml) | 147 (75, 277) | 719 (592, 866) |  |
| Female (%) | 259 (36) | 292 (52) | 1x10<sup>-8</sup>* |
| Age of Diagnosis (years) | 19.4 (12.9, 29.7) | 27.1 (17.5, 38.8) | 3x10<sup>-16</sup>* |
| Duration of Diabetes (weeks) | 11 (6, 18) | 10 (6, 17.5) | 0.43 |
| Hospital Admission (%) | 531 (75) | 396 (71) | 0.15 |
| DKA (%) | 303 (43) | 223 (41) | 0.40 |
| Polyuria (%) | 666 (95) | 534 (97) | 0.26 |
| Weight Loss (%) | 597 (85) | 479 (88) | 0.24 |
| HbA1c (mmol/mol) | 81 (57, 106) | 87 (64, 110) | 0.01 |
| BMI, kg/m<sup>2</sup> | 23.3 (20.9, 26.2) | 23.7 (21.5, 26.7) | 0.02 |
| Parent with Diabetes (%) | 92 (13) | 112 (20) | 6x10<sup>-4</sup>* |
| Other autoimmune condition (%) | 39 (5) | 71 (13) | 5x10<sup>-6</sup>* |
| T1D-GRS | 0.275 (0.256, 0.293) | 0.274 (0.253, 0.292) | 0.58 |
| HLA-DR3-DQ2 (%) | 359 (50) | 328 (59) | 3x10<sup>-3</sup> |
| HLA-DR4-DQ8 (%) | 382 (54) | 273 (49) | 0.10 |
| Number of positive autoantibodies |  |  | 0.43 |
| One (%) | 179 (25) | 148 (27) |  |
| Two (%) | 201 (28) | 170 (31) |  |
| Three (%) | 331 (47) | 239 (43) |  |
| IA-2A (%) | 454 (64) | 345 (62) | 0.48 |
| IA-2A level (DK U/ml)<sup>#</sup> | 255 (125, 340) | 254 (95, 339) | 0.59 |
| ZnT8A (%) | 409 (58) | 303 (54) | 0.27 |
| ZnT8A level (AU/ml)<sup>#</sup> | 36 (11, 78) | 43 (13, 86) | 0.13 |

We used a second independent replication cohort of 427 participants with clinically diagnosed type 1 diabetes (age at diagnosis range 17–81 years) and positivity to any of the three islet autoantibodies (GADA, IA-2A and ZnT8A). The participants were part of the UK-wide StartRight study (11). All were recruited <12 months from diagnosis, insulin-treated from diagnosis and self-reported to be of white European ethnicity. The overall cohort characteristics is provided in ESM Table 1.

### Islet autoantibody measurement

#### ADDRESS-2 study

The islet autoantibodies (GADA, IA-2A and ZnT8A) were measured using established radiobinding assays by the Diabetes and Metabolism group at the University of Bristol (Bristol, U.K.) (12; 13) at a median of 10.1 weeks diabetes duration. Results for GADA and IA-2A are expressed in digestive and kidney units/mL (DK units/mL) units/mL or arbitrary units (AU/mL) for ZnT8A calculated from standard curves consisting of diluted patient sera in antibody-negative sera from healthy donors. Positive thresholds were set at 97.5^th^ percentile of 974 control samples for GADA (≥33 DK U/ml), the 98^th^ percentile of 500 control samples for IA-2A (≥1.4 DK U/ml) and the 97.5^th^ percentile of 523 healthy school children for ZnT8A (≥1.8 AU/ml) (14). The laboratory participates in the Islet Autoantibody Standardisation Program (IASP) (ESM Table 2).

**Table 2:**
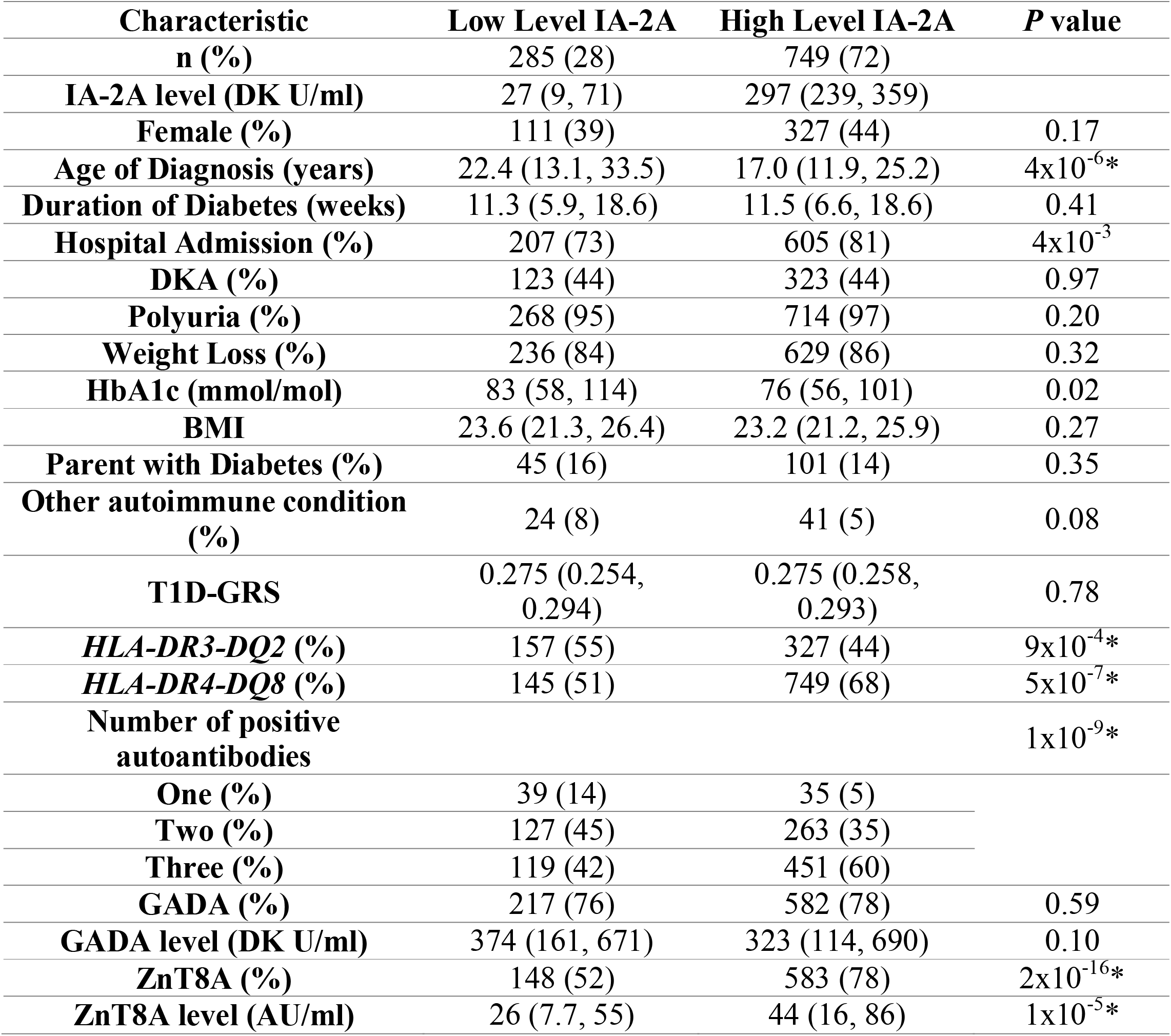
Comparison of clinical characteristics at diagnosis between high and lowIA-2A level groups for positive IA-2A type 1 diabetes cases. Bimodal IA-2A level distribution was divided into low and high level groups using the nadir between the mode at 130 DK U/ml. ^#^ only for people who were positive for that antibody * indicates a *P* value lower than threshold the *P* value for multiple comparisons (0.05/19 =0.0026)

| Characteristic | Low Level IA-2A | High Level IA-2A | P value |
| --- | --- | --- | --- |
| n (%) | 285 (28) | 749 (72) |  |
| IA-2A level (DK U/ml) | 27 (9, 71) | 297 (239, 359) |  |
| Female (%) | 111 (39) | 327 (44) | 0.17 |
| Age of Diagnosis (years) | 22.4 (13.1, 33.5) | 17.0 (11.9, 25.2) | 4x10<sup>-6</sup>* |
| Duration of Diabetes (weeks) | 11.3 (5.9, 18.6) | 11.5 (6.6, 18.6) | 0.41 |
| Hospital Admission (%) | 207 (73) | 605 (81) | 4x10<sup>-3</sup> |
| DKA (%) | 123 (44) | 323 (44) | 0.97 |
| Polyuria (%) | 268 (95) | 714 (97) | 0.20 |
| Weight Loss (%) | 236 (84) | 629 (86) | 0.32 |
| HbA1c (mmol/mol) | 83 (58, 114) | 76 (56, 101) | 0.02 |
| BMI | 23.6 (21.3, 26.4) | 23.2 (21.2, 25.9) | 0.27 |
| Parent with Diabetes (%) | 45 (16) | 101 (14) | 0.35 |
| Other autoimmune condition (%) | 24 (8) | 41 (5) | 0.08 |
| T1D-GRS | 0.275 (0.254, 0.294) | 0.275 (0.258, 0.293) | 0.78 |
| HLA-DR3-DQ2 (%) | 157 (55) | 327 (44) | 9x10<sup>-4</sup>* |
| HLA-DR4-DQ8 (%) | 145 (51) | 749 (68) | 5x10<sup>-7</sup>* |
| Number of positive autoantibodies |  |  | 1x10<sup>-9</sup>* |
| One (%) | 39 (14) | 35 (5) |  |
| Two (%) | 127 (45) | 263 (35) |  |
| Three (%) | 119 (42) | 451 (60) |  |
| GADA (%) | 217 (76) | 582 (78) | 0.59 |
| GADA level (DK U/ml) | 374 (161, 671) | 323 (114, 690) | 0.10 |
| ZnT8A (%) | 148 (52) | 583 (78) | 2x10<sup>-16</sup>* |
| ZnT8A level (AU/ml) | 26 (7.7, 55) | 44 (16, 86) | 1x10<sup>-5</sup>* |

#### StartRight study

GADA, IA-2A and ZnT8A islet autoantibodies were measured using ELISA assays (RSR Limited, Cardiff, U.K.) on a Dynex DS2 automated ELISA system (Launch Diagnostics, Longfield, U.K.), at a median of 14.9 weeks diabetes duration by the Academic Department of Blood Sciences, Royal Devon and Exeter Hospital (Exeter, U.K.) (15). Positive thresholds were set at the 97.5^th^ percentile of 1559 non-diabetic control subjects (GAD ≥ 11 WHO (World Health Organization) U/mL, IA-2 ≥ 7.5 WHO U/mL, ZnT8 age ≥ 30 years ≥ 10 U/mL, ZnT8 age < 30 years ≥ 65 U/mL). Upper reporting limits for GADA, IA-2A and ZnT8A were 2000 WHO U/mL, 4000 WHO U/mL and 2000 U/mL respectively. The laboratory also participates in IASP (ESM Table 2) (16).

### Type 1 diabetes genetic risk score and HLA genotypes

We generated weighted T1D-GRS from 30 common type 1 diabetes genetic variants (single nucleotide polymorphisms [SNPs]) for HLA and non-HLA loci as described in our previous papers (17). *HLA DR3-DQ2* and *HLA-DR4-DQ8* were imputed from two SNPs as described in our previous paper (15).

### Statistical Analysis

We used histograms to assess the distribution of islet autoantibody levels in those positive for that autoantibody. Autoantibody levels with bimodal distributions were split into high or low level categories using the nadir (lowest point between distributions). We also used a normal mixture model analysis and likelihood ratio tests to assess whether uni-modal, bi-modal or multi-modal distributions were best supported by the data. This analysis was performed on log-transformed autoantibody level data so that it was better approximated by a mixture of continuous, symmetric normal distributions. This was implemented using the mixtools package in R (18).

We performed Mann-Whitney tests to compare the continuous variables and Pearson chi-square tests were used to compare categorical variables between autoantibody level categories. All statistical analysis was carried out using Stata/SE 16.0 (StataCorp, College Station, TX) unless otherwise stated.

## Results

### GAD and IA-2 but not ZnT8 autoantibody levels exhibit a bimodal distribution at diagnosis in type 1 diabetes

We first assessed the distribution of GADA level in GADA positive type 1 diabetes people (n=1268), IA-2A level in IA-2A positive type 1 diabetes people (n=1034), and ZnT8A level in ZnT8A positive type 1 diabetes people (n=906). The distribution of the GADA and IA-2A levels showed two peaks consistent with a bimodal distribution (Fig. 1A & 1B). ZnT8A level showed a single peak with right-skewed distribution (Fig. 1C). Bimodality of GADA and IA-2A levels was also confirmed using a mixture model analysis (ESM Fig. 1A & 1B). Specifically, we analysed IA-2A and GADA levels on the log scale and used a likelihood ratio test (LRT) on 3 degrees of freedom to compare the log-likelihood of a one-component normal distribution with two parameters (one mean and one variance) versus that of a two-component, five parameter normal mixture (two means, two variances and a weight determining the relative proportion of each component). Under the null hypothesis that the single normal distribution was the true data generating model, twice the difference in log-likelihoods between the one-component and two-component models (referred to as the LRT) follows a chi-squared distribution on 5-2 = 3 degrees of freedom. Both analyses yielded overwhelming evidence in favour of the two-component model (LRT_IA-2A_ = 1129, p<2×10^−16^, LRT_GADA_= 345, p<2×10^−16^). We used the nadir value between the two peaks to divide the bimodal distribution of the autoantibody levels into two groups (low vs. high levels) for the subsequent analysis. The nadir value for GADA levels was 452 DK U/ml. All participants with a GADA level lower than this value were grouped into a low level GADA group (mean level 180.3, SD +/-118.4, 711/1268 (56%)) and the participants with a GADA level above or equal this value were grouped into a high level GADA group (760.9, +/-229.3, 557/1268 (44%)). Similarly, the nadir value of 130 DK U/ml between the two peaks of IA-2A levels divided people into a low level IA-2A group (mean level 41.2, SD+/-37.4, 285/1034 (28%)) and a high level IA-2A group (301.0, +/-88.3, 749/1034 (72%)).

**Figure 1:**
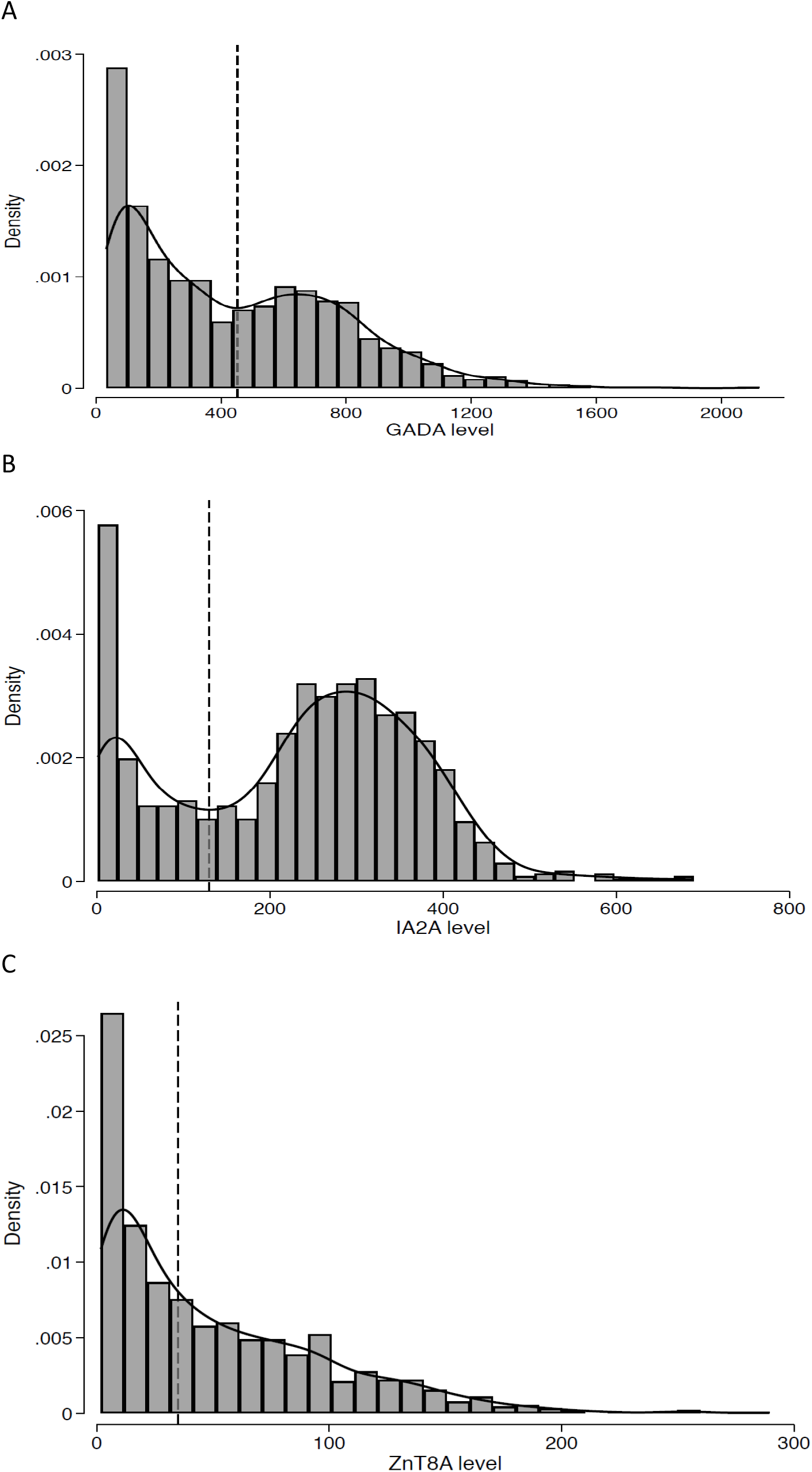
Histograms with kernel density curves showing the distribution of islet autoantibody levels in patients with type 1 diabetes at diagnosis. A) Histogram of GADA level at diagnosis measured using radiobinding assay for type 1 diabetes cases who were positive for GADA (n=1,268). GADA level exhibits a bimodal distribution. The nadir value of 452 DK U/ml between the two mode is highlighted with black dashed line and used to defined high GADA level group (≥452 DK U/ml) and low level group (<452 DK U/ml). B) Histogram of IA-2A level at diagnosis measured using radiobinding assay for type 1 diabetes cases who were positive for IA-2A (n=1,034). IA-2A level exhibits a bimodal distribution. The nadir value of 130 DK U/ml between the two mode is highlighted with black dashed line and used to defined high IA-2A level group (≥130 DK U/ml) and low level group (<130 DK U/ml). C) Histogram of ZnT8A levels at diagnosis measured using radiobinding assay for type 1 diabetes cases who were positive for ZnT8 (n=906) show a right skewed distribution. Median value of the distribution (35.4 AU/ml) is highlighted with black dashed lines and used to define high level ZnT8A (≥35.4 AU/ml) and low level (<35.4 AU/ml) groups.

### Higher GADA levels were associated with later age at diagnosis of type 1 diabetes, female sex, and *HLA-DR3-DQ2*

To assess the association of bimodal GADA levels to clinical features at diagnosis, we compared the clinical features between the people with low level (lower mode) and high level (higher mode) GADA as defined above (Table 1). Those in the high level GADA group were diagnosed later compared to the low GADA level group (median 27.1 y [IQR 17.5-38.8] vs. 19.4 y [12.9-29.7], *P*=3×10^−16^) (Fig. 2) (Table 1). They were more likely to be female (52% vs. 36%, *P*=1×10^−8^), and more likely to have other autoimmune diseases (13% vs. 5%, *P*=5×10^−6^) compared to low level GADA group (Table 1). They had modest enrichment for *HLA DR3-DQ2* (59% vs. 50%, *P*=3×10^−3^) but had similar T1D-GRS (median 0.275 [IQR 0.256-0.293] vs. 0.274 [0.253-0.292], *P*=0.53) based on 30 T1D associated common variants (19). The presentation characteristics (DKA, weight loss, polyuria, HbA1c and BMI), the number of other islet autoantibodies and other islet autoantibody levels were similar between the two GADA level groups.

**Figure 2:**
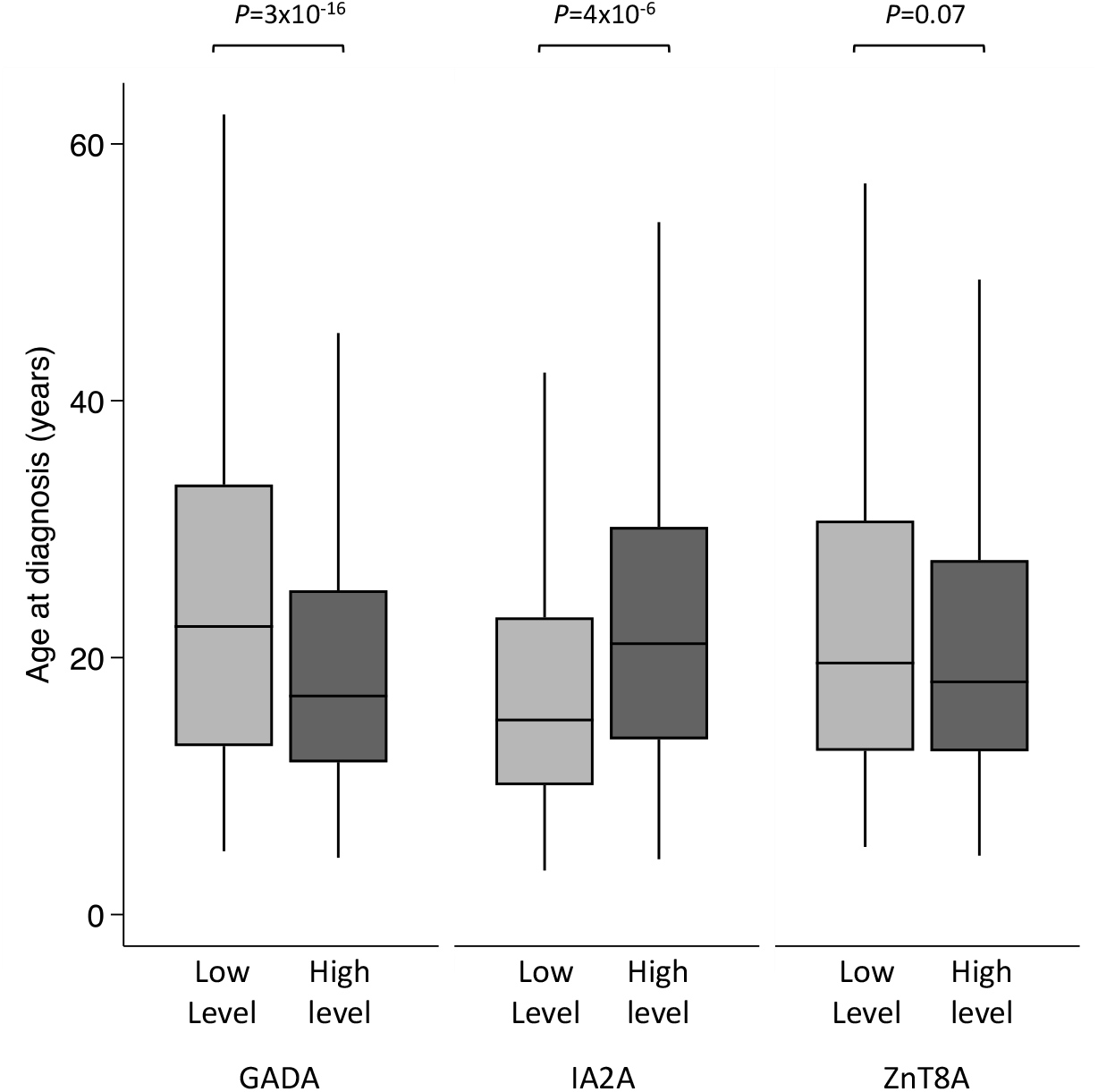
Box plot showing age of diagnosis of type 1 diabetes in high and low level groups for GADA, IA-2A and ZnT8A. The nadir value between the two modes of GADA level (452 DK U/mL) and IA-2A level (130 DK U/mL) distribution at diagnosis of type 1 diabetes who were positive for respective autoantibodies was used to define high and low level categories. There were 711/1,268 and 557/1,268 cases in low and high level GADA groups and 285/1,034 and 749/1,034 cases in low and high level IA-2A groups. The median value of ZnT8A level (35.4 AU/ml) was used for defining low and high level groups (n=453 each). Median age of diagnosis was higher for the high level GADA group (*P*=3×10^−16^), lower for the higher level IA-2A (*P*=4×10^−6^) and similar between ZnT8A level categories (*P*=0.07).

### Higher IA-2A levels were associated with earlier age at diagnosis of type 1 diabetes and *HLA DR4-DQ8* and ZnT8A positivity

We next compared the clinical features of low and high IA-2A level groups to assess association of bimodal IA-2A level distribution to clinical features at diagnosis. (Table 2). Contrary to GADA, those in the higher level IA-2A group were diagnosed earlier compared to the lower IA-2A level group (median 17.0 years [IQR 11.9-25.2 years] vs. 22.4 years [13.1-33.5 years], *P*=4×10^−6^) (Fig. 2) (Table 2). They were more likely to be multiple autoantibody positive (60% vs. 42% with three autoantibodies, *P*=1×10^−9^), positive for ZnT8A (78% vs. 52%, *P*=2×10^−16^) and more likely to have higher ZnT8A levels (median level 44 AU/ml [IQR 16-86] vs. 26 [7.7-55]) (*P* =1×10^−5^). Those with higher IA-2A levels were more likely to have *HLA DR4-DQ8* (68% vs. 51%, *P*=5×10^−7^). The presentation characteristics were similar between IA-2A level groups (Sex, DKA, weight loss, polyuria, HbA1c and BMI, parent with diabetes) (Table 2).

### ZnT8A level at diagnosis was not associated with age at diagnosis of type 1 diabetes

To assess the association of ZnT8A level to clinical features at diagnosis, we divided 906 T1D cases who were positive for ZnT8A by the median value of the distribution (35.4 AU/ml) due to lack of clear bimodal distribution. No statistically or clinically significant relationship was found between ZnT8A level and age at diagnosis (19.0 years [12.3-30.1] vs. 17.4 [12.2, 26.4], p=0.07) (Fig. 2) or *HLA DR3-DQ2* or *DR4-DQ8*. Both groups also exhibited similar presentation characteristics (DKA, weight loss, polyuria, HbA1c and BMI). However, those with higher level ZnT8A were more likely to be multiple autoantibody positive (71% vs. 55% with three autoantibodies) and more likely to be positive for IA-2A (86% vs. 76% *P*=1.5×10^−4^) (ESM Table 3) and GADA (83% vs. 74% *P*=0.002). Similar results for the lack of association of ZnT8A level to age at diagnosis were observed with linear regression analysis of log ZnT8A level to age at diagnosis (β=-0.57, 95% CI −1.2,0.08, *P*=0.09).

### Bimodal distributions of GAD and IA-2 islet autoantibody levels were also observed in second independent cohort

To replicate our results with different assay and different cohort, we analysed GADA, IA-2A and ZnT8A levels in 427 patients with type 1 diabetes at diagnosis from the StartRight study where islet autoantibody levels were measured using ELISA assays; another commonly used assay for islet autoantibody measurement.

Similar to our primary cohort, GADA and IA-2A levels showed a bimodal distribution in this replication cohort, with ZnT8A showing one peak with right skewed distribution (ESM Figure 2). The shape of the distribution is different in this cohort due to the clinical laboratory conducting the ELISAs not reporting results that are outside the standard curve leading to truncation at higher levels.

Using the same method as our primary cohort, we divided the GADA and IA-2A bimodal distributions into high and low autoantibody level groups using the nadir between the peaks (ESM Figure 2). Similarly to our primary analysis, those in the high level GADA group were diagnosed later compared to the low GADA level group (41 years [IQR 32-53 years] vs. 30 years [IQR 24-38 years], *P*=7×10^−13^), were more likely to be female (61% vs. 42%, *P*=2×10^−4^) and were more likely to have a concurrent autoimmune disease (22% vs. 11%, *P*=2×10^−3^) (ESM table 4). There were no differences in presentation characteristics, parent with diabetes, or C-peptide (ESM Table 4). The participants with higher IA-2A levels were younger (median 29 years vs. 33 years) compared to the ones with lower IA-2A levels, as our primary cohort, but this difference was not statistically significant (ESM Table 5).

## Discussion

Our study shows that GADA and IA-2A level at diagnosis of type 1 diabetes show clear bimodal distributions. Dichotomising levels into high or low groups according to the observed modes exhibits strong associations with age at diagnosis of type 1 diabetes but not with severity of type 1 diabetes at diagnosis.

There have been multiple studies of islet autoantibody levels in at-risk populations for type 1 diabetes (1; 7-9) and within type 1 diabetes populations but none of the studies to our knowledge have suggested that GADA and IA-2A have a bimodal distribution of level. However, the bimodality of GADA levels but not IA-2A levels has been reported in people with latent autoimmune diabetes (LADA) (20; 21). We were able to show the bimodality of levels using a second independent cohort of people with type 1 diabetes as well as using a second method for measuring islet autoantibodies. These data suggest that the bimodality of GADA and IA-2A levels is likely to be a true biological phenomenon, rather than an assay artefact, that is observed at diagnosis of type 1 diabetes.

The underlying biology of bimodality of GADA and IA-2A levels is not known and will need further investigation. We observed an enrichment of *HLA DR3-DQ2* and *HLA DR4-DQ8* in people with higher GADA levels and IA-2A levels respectively. Both these associations are well-described with the positivity of the respective autoantibodies but not with the autoantibody levels (22; 23). The difference in HLA susceptibility suggests a role for humoral immunity and antigen recognition as one of the factors underlying bimodality. However, the association with HLA was modest in our study and the overall genetic risk score was similar with high and low autoantibody level groups suggesting that there are additional factors which are responsible for the observed bimodality. Previous studies have shown that high GADA levels are correlated with higher affinity autoantibodies, the central and c-terminal epitopes, and multiple autoantibodies positivity (24). We did not observe the association of high GADA level with multiple autoantibodies, but we did observe the association of higher IA-2A level with multiple autoantibodies. These data suggest that affinity, the difference in epitopes, and the presence of other autoantibodies may also contribute towards bimodality.

GADA and IA-2A levels are associated with age at onset but in opposite directions. The participants who had high GADA levels were nearly 7 years older at diagnosis compared to the ones with low level GADA. This was replicated in a second independent cohort of adults with type 1 diabetes. Contrary to GADA, participants with high IA-2A levels were 5.4 years younger at diagnosis compared to those with low IA-2A levels. Interestingly, we note that both GADA and IA-2A levels but not ZnT8A level follows the same association with age as positivity of these autoantibodies (14; 25; 26). It has been shown that the pattern of positive autoantibody changes with age. The older-onset type 1 diabetes have a higher proportion of GADA whereas childhood-onset diabetes has a higher proportion of IA-2A, IAA and ZnT8A. The underlying reason for this observation is not clear. However, it is not due to change in overall humoral immunity due to age as we see the opposite effect of autoantibody levels on age with different autoantibodies. In contrast to our findings in type 1 diabetes, studies of LADA have shown that higher GADA level is associated with early age of onset. This may relate to the very different populations studied and the relationship between age and prior prevalence of autoimmune diabetes which, as recently suggested, may markedly alter antibody false positive rates in populations of apparent type 2 diabetes (21; 26). Studies of the at-risk population have shown higher IA-2A level is associated with earlier age of onset of type 1 diabetes supporting our observation of an inverse relationship between IA-2A level and age (27; 28). In contrast, studies of at-risk individuals have not reported an association of GADA level with the age of onset of type 1 diabetes (1; 28). The lack of inclusion of adults and focus on children in these studies may explain the lack of association.

We show that despite greatly different autoantibody levels, autoantibody level at diagnosis was not strongly associated with severity of presentation of type 1 diabetes as reflected by symptoms of hyperglycaemia, proportions with DKA, osmotic symptoms and weight loss, as well as C-peptide and HbA1c at diagnosis. These results contrast previous studies in LADA where high levels of GADA were associated with lower BMI, C-peptide and higher HbA1c (21). In contrast to LADA, higher levels of ICA but not GADA and IA-2A at diagnosis of type 1 diabetes was associated with C-peptide <50 pmol at 2 years (29). In a recent cross-sectional study of patients with type 1 diabetes; high GADA levels, but not IA-2A and ZnT8A levels, were individually associated with lower c-peptide (<2.5 pmol). However, in this study islet autoantibodies were measured at median 7.5 years after diagnosis and association of GADA levels were only seen at 2.1 years (30). Further longitudinal studies are needed to assess the association of bimodality of autoantibody level at diagnosis with beta-cell function over time in type 1 diabetes.

Our findings have important implications for the prediction, treatment and prognosis of type 1 diabetes. It is well known that type 1 diabetes is a heterogeneous disease with heterogeneity in islet autoantibodies, beta-cell function, genetics as well as response to immunomodulatory therapy (31). The research to date mainly focuses on the positivity of autoantibodies rather than autoantibody levels in type 1 diabetes. We believe that autoantibody levels showing a bimodality for GADA and IA-2A is an important consideration in understanding the heterogeneity of type 1 diabetes. It is well known that immunomodulatory therapy has a variable response on beta-cell function in clinical trials (32). Currently, the reason for this variable response is not entirely known but is proposed to be due to variation in T cell response (33; 34). The bimodality of the levels may represent a surrogate marker of a specific immune response and identify the subgroup of individuals with differential response to immunomodulatory therapy but this needs testing in further studies. This, along with the association of GADA level with c-peptide in a recent study, provides an exciting possibility of a stratified approach to type 1 diabetes treatment and prognosis which is currently lacking (30). Our findings also make a strong case to assess the usefulness of the bimodality of GADA and IA-2A levels in the prediction models of progression of diabetes in at-risk population in addition to autoantibody positivity.

Our autoantibody levels were assessed using radiobinding assays in our primary cohort. We did not measure autoantibody levels by ELISA on the same cohort to validate our findings using this more-commonly used assay in routine clinical laboratories. However, we observed similar results with ELISA assay in the second cohort; both laboratories participate in IASP and a high correlation of levels between the ELISA and radiobinding assay (35; 36) meaning that our findings are applicable to levels measured by both methods. Although a bimodal distribution of GADA and IA-2A was observed using both assay methods, the shape of the distributions does look different between assays. We believe this is due to the clinical laboratory conducting the ELISAs not reporting results that are outside the standard curve causing truncation at both ends in comparison the RBA laboratory which reports extrapolated results.

Our study is limited to white-European populations, due to lack of validity of T1D GRS in non-European ethnicity and further studies are needed in non-white ethnicity. Also, we only had c-peptide information at diagnosis and only in one of the study cohorts, therefore we are limited in our ability to assess the impact of autoantibody level distribution on beta cell function. We did not study insulin autoantibodies (IAA) as this would not be appropriate due to the nature of our cohorts being recruited weeks after diagnosis of type 1 diabetes and commencement of insulin therapy.

In conclusion, we show that GADA and IA-2A levels exhibit a bimodal distribution at diagnosis of type 1 diabetes, which is biologically important to the understanding of the heterogeneity of type 1 diabetes and opens the exciting possibility of further research to assess its implication on prediction, treatment and prognosis of type 1 diabetes.

## Supporting information

Electronic Supplemental Material

## Data Availability

Data may be available upon reasonable request

## Article Information

## Acknowledgements

The authors acknowledge the staff of the Diabetes and Metabolism Group (University of Bristol) and of the Academic Department of Biochemistry (Royal Devon and Exeter NHS Trust) for their role in collecting the autoantibody data. K.A.P is the guarantor of this work and, as such, has full access to all the data in the study and takes responsibility for the integrity of the data and the accuracy of the data analysis. Finally, the authors would like to thank all the participants of the ADDRESS-2 and StartRight studies.

## Author Contributions

S.L.G assayed the Address-2 autoantibody samples at Bristol, researched the data and wrote the manuscript. S.L.G, K.A.P and J.B researched the data and wrote the manuscript. A.G.J and T.J.McD helped plan the research, contributed to the discussion and reviewed/edited the manuscript. B.M.S and T.J.McK contributed to data analysis and reviewed/edited the manuscript. A.G.J set up the study from which the replication cohort was selected. H.C.W, A.K, S.M, N.S.O and D.G.J. set up the ADDRESS-2 study from which the original cohort was selected, collected the clinical data and reviewed/edited the manuscript. This work, in part, has been presented as a poster at the nPOD conference (2021) and the Diabetes UK Professional Conference (2021).

