## Supplementary material for "Islet autoantibody level distributions in type 1 diabetes and their association with genetic and clinical characteristics": Electronic Supplemental Material

| Characteristic | ADDRESS-2 | StartRight |
| --- | --- | --- |
| n | 1,536 | 427 |
| Female (%) | 648 (42) | 208 (49) |
| Age of Diagnosis (years) | 20.8 (13.1, 31.5) | 34 (26, 47) |
| Duration of Diabetes at Antibody testing (weeks) | 10.1 (6.0, 16.9) | 14.9 (6.1, 31.7) |
| Hospital Admission (%) | 1,158 (76) | 253 (59) |
| DKA (%) | 636 (42) | 149 (35) |
| Polyuria (%) | 1,458 (96) | 389 (91) |
| Weight Loss (%) | 1,296 (86) | 368 (86) |
| HbA1c (mmol/mol) | 81 (59, 107) | 64 (50, 87) |
| BMI (kg/m^2^) | 23.4 (21.2, 56.2) | 24.5 (21.8, 27.2) |
| C-Peptide (picomol/L) | - | 458 (273, 673) |
| Parent with Diabetes (%) | 239 (16) | 85 (20) |
| Other autoimmune condition (%) | 116 (8) | 64 (15) |
| T1D-GRS | 0.275 (0.256, 0.293) | - |
| *HLA-DR3-DQ2* (%) | 800 (52) | - |
| *HLA-DR4-DQ8* (%) | 839 (55) | - |
| Number of positive autoantibodies |  |  |
| *One (%)* | 434 (28) | 148 (35) |
| *Two (%)* | 532 (35) | 124 (29) |
| *Three (%)* | 570 (37) | 155 (36) |
| GADA positive (%) | 1,268 (83) | 383 (90) |
| IA-2A positive (%) | 1,034 (67) | 227 (53) |
| ZnT8A positive (%) | 906 (59) | 251 (59) |

**ESM Table 1: Clinical characteristics for the study cohorts. ‘-‘** indicates unavailable data for this cohort. Values expressed as median (interquartile range) unless stated.

| Method | Antibody | %Sensitivity | %Specificity | IASP Workshop |
| --- | --- | --- | --- | --- |
| RBA | GADA | 74 | 96.7 | 2015 |
| RBA | IA-2A | 72 | 100 | 2015 |
| RBA | ZnT8RA | 60 | 100 | 2015 |
| RBA | ZnT8WA | 46 | 100 | 2015 |
| RSR ELISA | GADA | 74.0 | 98.9 | 2020 |
| RSR ELISA | IA-2A | 72.0 | 98.9 | 2020 |
| RSR ELISA | ZnT8A | 74.0 | 98.9 | 2020 |

**ESM Table 2: IASP Workshop performance for each islet autoantibody assay.** The radiobinding assays were all conducted centrally in Bristol (U.K.) by the Diabetes and Metabolism Group. The RSR ELISA assays were all conducted centrally by the Academic Department of Clinical Biochemistry (Royal Devon and Exeter NHS Trust) in Exeter (U.K.). RBA: Radiobinding immunoassay; ELISA: Enzyme-linked immunosorbent assay.

| Characteristic | Low Level ZnT8A | High Level ZnT8A | *P* value |
| --- | --- | --- | --- |
| n (%) | 453 (50) | 453 (50) |  |
| ZnT8A level (AU/ml) | 11.0 (5.0, 20.3) | 78.4 (54.1, 111.8) |  |
| Female (%) | 182 (40) | 196 (43) | 0.44 |
| Age of Diagnosis (years) | 19.0 (12.3, 30.1) | 17.4 (12.2, 26.4) | 0.07 |
| Duration of Diabetes (weeks) | 10.3 (5.9, 17.0) | 10.1 (6.1, 16.9) | 0.97 |
| Hospital Admission (%) | 368 (81) | 351 (78) | 0.20 |
| DKA (%) | 210 (47) | 184 (41) | 0.09 |
| Polyuria (%) | 433 (97) | 438 (98) | 0.29 |
| Weight Loss (%) | 387 (87) | 379 (85) | 0.44 |
| HbA1c (mmol/mol) | 80 (58, 113) | 78 (58, 102) | 0.38 |
| BMI | 23.3 (21.0, 25.5) | 23.3 (21.3, 26.5) | 0.23 |
| Parent with Diabetes (%) | 67 (15) | 56 (13) | 0.31 |
| Other autoimmune condition (%) | 31 (7) | 35 (8) | 0.61 |
| T1D-GRS | 0.274 (0.256, 0.294) | 0.276 (0.258, 0.293) | 0.61 |
| *HLA-DR3-DQ2* (%) | 222 (49) | 243 (54) | 0.16 |
| *HLA-DR4-DQ8* (%) | 271 (60) | 254 (56) | 0.25 |
| Number of positive autoantibodies |  |  | 4x10^-6^* |
| *One (%)* | 23 (5) | 10 (2) |  |
| *Two (%)* | 180 (40) | 123 (27) |  |
| *Three (%)* | 250 (55) | 320 (71) |  |
| IA-2A (%) | 343 (76) | 388 (86) | 1.5x10^-4^* |
| IA-2A level (DK U/ml)^#^ | 248 (124, 326) | 286 (214, 358) | 7x10^-5^* |
| GADA (%) | 337 (74) | 375 (83) | 2x10^-3^* |
| GADA level (DK U/ml) ^#^ | 298 (111, 675) | 361 (141, 722) | 0.05 |

**ESM Table 3: Comparison of clinical characteristics between high and low level ZnT8A groups for positive ZnT8A type 1 diabetes cases.** ZnT8A level distribution was divided into two groups using the median of the distribution at 35.4 AU/ml. Values expressed as median (interquartile range) unless stated. ^#^ only for people who were positive for that antibody * indicates a p value lower than threshold the p value for multiple comparisons (0.05/19 =0.0026)

| Characteristic | Low Level GADA | High Level GADA | *P* value |
| --- | --- | --- | --- |
| n (%) | 205 (54) | 178 (46) |  |
| GADA level (U/ml) | 93.6 (44.3, 319.6) | 2001 (1849, 2001) |  |
| Female (%) | 119 (42) | 70 (61) | 2x10^-4^* |
| Age of Diagnosis (years) | 30 (24, 38) | 41 (32, 53) | 7x10^-13^* |
| Duration of Diabetes (weeks) | 14.4 (5.7, 31.6) | 14.9 (7, 30.7) | 0.53 |
| Hospital Admission (%) | 128 (63) | 101 (57) | 0.23 |
| DKA (%) | 72 (35) | 63 (35) | 0.98 |
| Polyuria (%) | 190 (93) | 159 (89) | 0.25 |
| Weight Loss (%) | 169 (82) | 161 (90) | 0.02 |
| HbA1c (mmol/mol) | 65 (50, 87) | 63.5 (50.5, 88) | 0.77 |
| BMI (kg/m^2^) | 24.2 (21.5, 26.8) | 24.8 (21.9, 27.5) | 0.18 |
| C-Peptide (picomol/L) | 473 (273, 683) | 435 (266, 642) | 0.62 |
| Parent with Diabetes (%) | 37 (18) | 40 (22) | 0.28 |
| Other autoimmune condition (%) | 22 (11) | 39 (22) | 2x10^-3^* |
| Number of positive autoantibodies |  |  | 0.94 |
| *One (%)* | 61 (30) | 54 (30) |  |
| *Two (%)* | 62 (30) | 51 (29) |  |
| *Three (%)* | 82 (40) | 73 (41) |  |
| IA-2A (%) | 106 (52) | 90 (51) | 0.82 |
| IA-2A level (DK U/ml)^#^ | 328 (84, 1900) | 245 (40, 1575) | 0.22 |
| ZnT8A (%) | 120 (59) | 107 (60) | 0.75 |
| ZnT8A level (AU/ml) ^#^ | 228 (93, 571) | 279 (88, 592) | 0.69 |

**ESM Table 4: Comparison of clinical characteristics between high and low level GADA groups for positive GADA type 1 diabetes cases in StartRight study.** Bimodal GADA level distribution was divided into two groups using the nadir between the two modes at 1022 WHO U/ml. Values expressed as median (interquartile range) unless stated. ^#^ only for people who were positive for that antibody; * indicates a p value lower than threshold the p value for multiple comparisons (0.05/17 =0.0029)

| Characteristic | Low Level  IA-2A | High Level  IA-2A | *P* value |
| --- | --- | --- | --- |
| n (%) | 185 (81) | 42 (19) |  |
| IA-2A level (U/ml) | 141.3 (38, 558.1) | 4001 (3999, 4001) |  |
| Female (%) | 183 (45) | 23 (55) | 0.25 |
| Age of Diagnosis (years) | 33 (25, 97) | 29 (22, 48) | 0.34 |
| Duration of Diabetes (weeks) | 14.9 (6.4, 34.6) | 11.6 (6.6, 16.7) | 0.04 |
| Hospital Admission (%) | 112 (61) | 42 (67) | 0.49 |
| DKA (%) | 68 (37) | 14 (33) | 0.66 |
| Polyuria (%) | 169 (91) | 40 (95) | 0.40 |
| Weight Loss (%) | 161 (87) | 31 (74) | 0.03 |
| HbA1c (mmol/mol) | 63 (49, 87) | 66 (57, 91) | 0.46 |
| BMI (kg/m^2^) | 24.6 (22.3, 27.0) | 25.4 (21.2, 29.2) | 0.49 |
| C-Peptide (picomol/L) | 461 (298, 638) | 454.5 (301, 756) | 0.67 |
| Parent with Diabetes (%) | 29 (16) | 8 (19) | 0.59 |
| Other autoimmune condition (%) | 37 (17) | 7 (13) | 0.46 |
| Number of positive autoantibodies |  |  | 0.97 |
| *One (%)* | 18 (8) | 4 (7) |  |
| *Two (%)* | 55 (25) | 14 (26) |  |
| *Three (%)* | 143 (66) | 36 (67) |  |
| GADA (%) | 159 (86) | 37 (88) | 0.71 |
| GADA level (DK U/ml) | 917 (97, 2001) | 439 (76, 2001) | 0.87 |
| ZnT8A (%) | 134 (72) | 32 (76) | 0.62 |
| ZnT8A level (AU/ml) | 579 (11, 612) | 533 9295, 1090) | 4x10^-3^* |

**ESM Table 5: Comparison of clinical characteristics between high and low level IA-2A groups for positive for IA-2A type 1 diabetes cases in StartRight study.** Bimodal IA-2A level distribution was divided into low and high level groups using the nadir between the modes at 2750 WHO U/ml. ^#^ only for people who were positive for that antibody. Values expressed as median (interquartile range) unless stated. *Indicates a p value lower than threshold the p value for multiple comparisons (0.05/17 =0.0029)

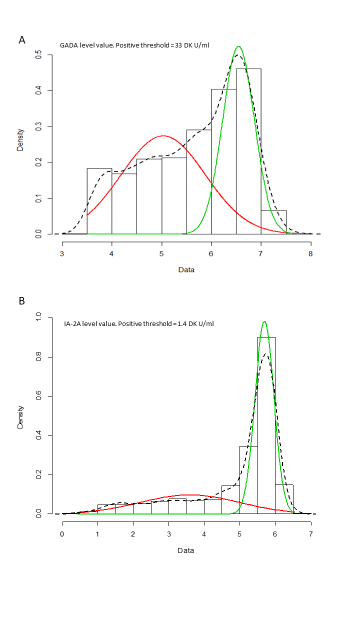

**ESM Figure 1: Log autoantibody level density plots showing the two best fitting normal densities for GADA and IA-2A level distributions.** A) Log GADA level density plot. Histogram and black dashed line show observed density. Red and green lines show the two best fitting normal densities which are used in the calculation of the likelihood ratio test. Likelihood ratio test: normal distribution versus two component mixture distribution: Log(GADA) ~ *N*(θ, σ^2^) vs. Log(GADA) ~ *w_1_N*(θ_1_, $\sigma_{1}^{2}$)+*w*_2_*N*(θ_2_, $\sigma_{2}^{2}$). LRT = 345 on 3 degrees of freedom, p-value = 0. B) Log IA-2A level density plot. Histogram and black dashed line show observed density. Red and green lines show the two best fitting normal densities which are used in the calculation of the likelihood ratio test**.** Likelihood ratio test: univariate normal distribution versus two component mixture distribution: Log(IA-2A) ~ *N*(θ, σ^2^) vs. Log(IA-2A) ~ *w*_1_*N*(θ_1_, σ^2^)+*w*_2_*N*(θ_2_, σ^2^). LRT = 1129 on 3 degrees of freedom, p-value = 0.

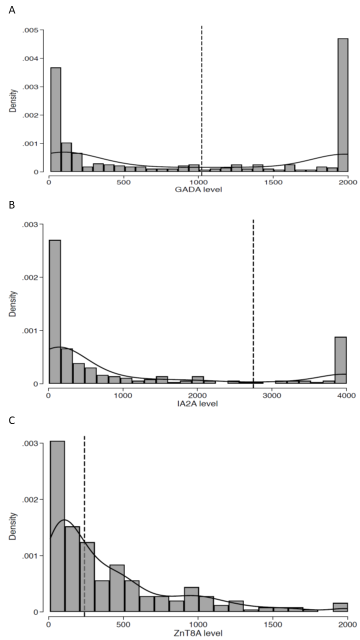

**ESM Figure 2: Histograms with kernel density curves showing the distribution of islet autoantibody levels at diagnosis measured using ELISA assay in patients with type 1 diabetes in the replication cohort (StartRight cohort).** A) Histogram of GADA level for type 1 diabetes cases who were positive for GADA (n=383) at diagnosis. GADA levels exhibit a bimodal distribution. The ELISA assay was calibrated to maximum value of 2000 WHO U/ml. The nadir value of 1022 WHO U/ml between the two modes is highlighted with black dashed line and used to defined high level group (≥1022 WHO U/ml) and low level group (<1022 WHO U/ml). B) Histogram of IA-2A level for type 1 diabetes cases who were positive for IA-2A (n=227) at diagnosis. IA-2A levels exhibit a bimodal distribution. The ELISA assay was calibrated to maximum value of 4000 WHO U/ml. The nadir value of 2750 WHO U/ml between the two modes is highlighted with black dashed line and used to defined high level group (≥2750 WHO U/ml) and low level group (<2750 WHO U/ml). C) Histogram of ZnT8A level for type 1 diabetes cases positive for ZnT8A (n=251) at diagnosis show a right skewed distribution. The ELISA assay was calibrated to maximum value of 2000 WHO U/ml. Median value of the distribution (240 AU/ml) is highlighted with black dashed lines.
